# Validating a previously untested ‘Intentions and Beliefs around Smoking’ sub-scale for inclusion in the published ‘Attitudes and Beliefs about Cardiovascular Disease (ABCD) Risk Questionnaire’ using a cross-sectional sample

**DOI:** 10.1101/2021.06.10.21258608

**Authors:** Mark Bowyer, Hamid Yimam Hassen

## Abstract

**Objectives:** To provide evidence of validity, reliability and generalisability of results obtained using the Attitudes and Beliefs about Cardiovascular Disease (ABCD) Risk Questionnaire with a sample of the English population surveyed within the ‘SPICES’ Horizon 2020 project (Nottingham study site), and to specifically evaluate the psychometric and factor properties of an as-yet untested 5 item sub-scale relating to smoking behaviours.

**Design and setting:** Community based cross-sectional study in Nottingham, UK.

**Participants:** 466 English adults fitting inclusion criteria (aged 18+, without known history of CVD, not pregnant, able to provide informed consent) were included in the study.

**Methods:** We re-validated the published ABCD questionnaire on a sample of the general population in Nottingham to confirm the psychometric properties. Furthermore, we introduced 5 items related to smoking which were dropped in the original study due to inadequate valid samples.

**Primary and secondary outcome measures:**

- Psychometric and factor performance of untested 5 item ‘smoking behaviours’ sub-scale
- Psychometric and factorial properties in combination with the remaining 18 items across 3 sub-scales

**Results:** Analyses of the data largely confirmed the validity, reliability, and factor structure of the original ABCD Risk Questionnaire. Sufficient participants in our study provided data against an additional five smoking related items to confirm their validity as a sub-scale and to advocate for their inclusion in future applications of the scale. EFA and CFA calculations support some minor changes to the remaining sub-scales which may further improve psychometric performance and therefore generalisability of the instrument.

**Conclusions:** An amended version of the ABCD Risk Questionnaire would provide public health researchers and practitioners with a brief, easy to use, reliable and valid survey tool. The amended tool may now assist public health practitioners and researchers to quickly survey patient or public intentions and beliefs around three key areas of individually modifiable risk (Physical Activity, Diet, and Smoking).

**Trial registration:** ISRCTN68334579 https://doi.org/10.1186/ISRCTN68334579

Heart health without a doctor: an implementation study of CVD prevention and behaviour change interventions in community settings

**Ethical approval:** Ethical approval for the ‘SPICES’ Nottingham study protocol (incorporating the ABCD Risk Questionnaire) was secured from the Nottingham Trent University College of Business, Law and Social Sciences on the 20^th^ February 2019. Participants were required to provide informed consent (Appendix 4).

**Article summary (Strengths and Limitations of this study):**

- Large sample (n=466) of English adults from the Nottingham UK population
- Sufficient case data to validate additional sub-scale related to attitudes and intentions of smokers
- Criterion validity not explored
- Full assessment of the utility of ABCD Risk Questionnaire in health promotion and CVD prevention not explored, further studies may be required to position the tool in clinical and public health practice.

**Original protocol:** (Appendix 3)

**Funding statement:** This work was supported by the European Commission Horizon 2020 Non-communicable diseases and the challenge of healthy ageing Grant agreement 733356 ‘SPICES’.

**Competing interests statement:** None declared

**Patient and public involvement:** Patients and/or the public were not involved in the design, or conduct, or reporting, or dissemination plans of this research.

**Patient consent for publication:** (data sharing agreement)

Not required (participant information and informed consent attached Appendix 4)

**Provenance and peer review:** Not commissioned.

**Data availability statement:** Data are available on reasonable request

**Author statement:** Mark Bowyer: Design of work, acquisition of data, analysis and interpretation of data, drafting and revising the paper, final approval, accountability for accuracy and integrity.

Hamid Hassen: Analysis and interpretation of data, drafting and interpretation of results, accountability for accuracy and integrity.

## INTRODUCTION

### Scientific Background and Rationale

In the UK, Cardiovascular Disease (CVD) is responsible for over 130,000 deaths per annum ^i^. CVD morbidity is also the biggest contributor to the inequalities in Healthy Life Expectancy between members of the wealthiest neighbourhoods and the most deprived^ii^. In 2009 the NHS Health Check^iii^ was established and more recently (2019) the CVD Prevent^iv^ initiative to implement ‘upstream’ interventions for the prevention of CVD morbidity. Both of these initiatives seek to improve early case-finding to prevent avoidable strokes and heart attacks. Both recognise the importance of supported lifestyle change in conjunction with drug therapies.

Lifestyle or behavioural change requires a degree of individual agency and commitment which drug therapies do not. Unhealthy lifestyle behaviours are linked to culture and habit, environment, emotions, and confidence which can all moderate an individual’s readiness to change and the commitment required to sustain those changes over time^v^. Understanding the attitudes and beliefs that people hold towards diet, exercise and smoking, as well as their perception of their own risk could assist primary care and public health professionals in providing relevant and effective behavioural advice and social prescribing options. To support evaluations of the NHS Health Check programme, in 2017 a questionnaire was developed to evaluate patients’ awareness of cardiovascular disease risk at University College London^vi^. This ABCD Risk Questionnaire attempts to provide a short survey drawing from the dominant theoretical models of behaviour change (Trans-Theoretical Model, Health Beliefs Model)^vii^, covering diet, smoking, exercise and alcohol behaviours, and incorporating a conceptual spread of perceived risk from immediate to lifetime.

### Specific Objectives

In this study we re-validated the tool on a sample of the general population in Nottingham to confirm the psychometric and factorial properties. Furthermore, we introduced 5 items related to smoking which were dropped in the original study due to inadequate case numbers.

To the best of our knowledge, this is the first study which has incorporated items relating to attitudes and intentions towards stopping smoking into the published version of the ABCD Risk Questionnaire and collected sufficient data to submit them to analysis of validity, reliability and factor structure.

In the original ABCD study, over the course of three stages of validity testing (content, face, reliability) items relating to alcohol use and smoking were rejected, leaving four final sub-scales: Knowledge of CVD Risks; Perceived Risk of Heart Attack/ Stroke; Perceived Benefits and Intentions to Change; and Healthy Eating Intentions. During Exploratory Factor Analysis (EFA) none of the items relating to alcohol use achieved strong enough loadings to be included in the final scale, and items related to smoking could not be included due to the high proportion of missing data in the experimental sample. The authors of the study note this limitation *‘the questionnaire does not encompass all aspects of CVD risk observed in the general population’ and that ‘future studies examining populations at increased CVD risk can look into incorporating smoking and alcohol into the ABCD Risk Questionnaire to learn about these individuals’ preconceptions and attendance of follow-up care’*^*viii*^.

### The present study

Nottingham is one of five global sites of the EU Horizon 2020 ‘SPICES’^ix^ CVD prevention implementation study which began in 2017. SPICES investigates contextual and health system barriers to the scaling up of successful behaviour change interventions for improved cardiovascular health in low, middle and high income European countries.

The SPICES Nottingham population survey carried out in 2019-20 utilised the ABCD Risk Questionnaire alongside the non-clinical INTERHEART CVD risk prediction instrument^x^. The SPICES study team chose to re-introduce 5 pre-written items relating to ‘Intentions and Readiness to Stop Smoking’ from the 65 item University College London (UCL) item pool into the questionnaire due to the high prevalence of smoking in the Nottingham population compared to England averages^xi^, and its importance as a CVD risk^xii^. This created a 31 item questionnaire.

In so doing, NTU researchers attempted to *‘replicate the factor analytic process on an independent, larger sample to confirm the generalisability of (the original) findings’ as requested by the authors of the original study. At the same time, we anticipated securing sufficient responses against the reintroduced 5 item ‘smoking’*^*xiii*^ sub-scale to analyse its reliability and validity as an integral part of future versions of the Questionnaire.

## METHODS

Incorporating the ABCD Risk Questionnaire into the SPICES Nottingham baseline survey provided cross-sectional study data across a broad sample of adult participants. The data-set generated was therefore suitable for psychometric validation of the original and modified versions of the ABCD questionnaire.

### Participants

Participants were recruited from across the Nottingham conurbation between April 2019 and March 2020 as part of the SPICES Nottingham^xiv^ baseline survey. A purposive sampling method was employed based on community engagement. This strategy had two components:

1. engagement of citizens in neighbourhoods through existing community groups, organisations and venues, and

2. engagement of employees in the workplace through large city-based employers.

Community groups were targeted on the basis of the demographic of their membership to ensure that neighbourhoods of differing mean household income, those who are not in employment or of working age, and those from different ethnicities were included. In this way 327 participants were recruited.

Employers were targeted on the basis of workforce size, and policies relating to workforce well-being. Nottingham City Council Adult Care teams and the Rolls-Royce plc Hucknall site both responded positively and between them provided 156 participants. NTU researchers administered the SPICES Nottingham baseline survey individually within the community or workplace setting and personalised feedback about CVD risks was provided confidentially once the survey had been completed.

### Materials

The SPICES baseline survey incorporated the ABCD risk questionnaire into a digitised survey instrument created in the Research Electronic Data Capture (REDCap) database system^xv^, a secure web application for building and managing online surveys and databases, and the online survey responses were uploaded automatically. No participant data was stored on local devices. Both the ABCD Risk Questionnaire and the non-laboratory INTERHEART questionnaire^xvi^ were included unchanged from their published versions apart from an additional 5 items pertaining to smoking behaviour.

The surveys were administered in the field by a team of trained researchers recruited from the NTU student body and directly supervised by the SPICES Nottingham coordinator. The surveys were accessed using dedicated tablet computers. Items were reproduced word for word and in the same sequence as the original ABCD Risk Questionnaire with the additional 5 smoking items inserted after all 26 original items.

### Validating the sample

The baseline survey dataset was extracted from REDCap for analysis. Sample was checked for representativeness of the Nottingham population across parameters of age, gender, household income and known rates of physical activity and smoking.

### Data analysis

We took the published 26-item ABCD Risk Questionnaire, introduced 5 further items relating to smoking behaviours, and administered it alongside a validated CVD risk assessment instrument (INTERHEART) to 486 individuals in Nottingham over a period of 12 months. Item, scale, and factor reliabilities were remeasured. Correlation was tested between and amongst ABCD sub-scale scores and selected INTERHEART variables, closely matching the methods applied in the original study (Appendix 6) and results were compared accordingly. After data cleansing, 466 valid cases were entered for analysis, four times the sample size of the original study.

Item and sub-scale reliabilities were tested using inter-item correlations, corrected item-total correlations and Cronbach’s Alpha.^xvii^ We performed an exploratory factor analysis (EFA)^xviii^to evaluate the dimensionality of items of the original and modified risk scale with and without the smoking items. The EFA was performed using the maximum likelihood extraction and varimax rotation method.^xix^ Sample and data adequacy was assessed using Kaiser-Meyer-Olkin (KMO) test and Bartlett’s test of sphericity^xx^ was performed to compare an observed correlation matrix to the identity matrix. The adequate number of factors was determined using a scree plot. To further test the consistency of factors, we tested using Confirmatory Factor Analysis (CFA). We evaluated the model fit of the CFA using; the X2 test, the Tucker-Lewis and Comparative Fit Indexes and the root mean square error of approximation (RMSEA)^xxi^. The analysis was performed using a free statistical software R version 4.0.2. UK postcodes were collected for all participants which allowed them to be sorted into income deciles using Office for National Statistics Index of Multiple Deprivation (IMD) public datasets, allowing correlations to be analysed. Case data from the ‘Knowledge’ sub-scale (8 items) were omitted from the analysis since they utilise a separate response format.

We used the STROBE cross sectional checklist when writing our report^xxii^.

## RESULTS

### Participants

Participation was voluntary, and self-selection may have been influenced by sensitivities around disclosure of health status and lifestyle habits forming a barrier to those with co-morbidities and socially ‘questionable’ behaviours (heavy smoking, high alcohol intake).

The sample cohort is strongly parametric, with a 49:51 percent gender split, normal distribution of age ranges (18-92), and a distribution of Socio-Economic Status (SES) which reflects known data about neighbourhood income in Nottingham. Nottingham is the 11^th^ most deprived district in England with higher unemployment, lower education and skills, and shorter life expectancy than the national averages^xxiii^. Using the Index of Multiple Deprivation a relative measure of deprivation across seven domains, Health and Disability s the domain on which the city does worst. Nevertheless, the mean INTERHEART predicted risk score for all 466 participants was 10.32 which closely matches the global reported mean for the instrument^xxiv^.

### Smoking sub-scale

The percentage of smokers in our sample was 15.5%. The five items in the smoking subscale are measured on the same four-point response scale as the 18 items submitted for Factor Analysis in the original published ABCD Risk Questionnaire (Strongly agree, agree, disagree, strongly disagree, and not applicable).

With the original 18 items this ‘Not Applicable’ response option was not used by any of the SPICES Nottingham study participants. By contrast, within their responses to the items in the ‘smoking’ subscale, ‘Not Applicable’ was the modal answer. Participants chose the ‘N/A’ response option whenever they reported being a non-smoker. This mirrors the behaviour of the original 110 NHS Health Check attendees who formed the pilot sample cohort for the original study, leaving an insufficient number of cases to assess validity and reliability of smoking sub-scale items. In the present study, 88 cases were found where participants reported smoking behaviours and this was sufficient to enter them into analysis.

Sub-scale Alpha values, Cronbach’s Alpha if item deleted calculated for all items, inter-item correlations and corrected item-total correlations were all calculated, mirroring the analysis reported in the original study.

Interitem correlations calculated for these five items produced a range between 0.654 and 0.834. All of these five ‘smoking’ items therefore correlate with one another more strongly than recommended (<.6) and were considered for rejection. However, we found each item to be qualitatively different, and that the differences were conceptually clear and well expressed in the item wording so that no participant could be expected to confuse one with any other, and they were retained.

Discrimination was confirmed using item-total correlations. These fell between the range 0.751 and 0.906 meaning that all five ‘smoking’ sub-scale items are comfortably above the standard cut-off for acceptability of 0.3.

EFA was carried out twice, firstly with the 88 confirmed smoking cases, and then again with all cases. The first operation ensured that factor loadings were not skewed by the lower number of cases reporting smoking behaviours, the second ensured that factor loadings for the remaining sub-scales where more case data was available were not skewed by outliers.

### Exploratory Factor Analysis

We conducted EFA on the original 18-item risk perception questionnaire and the modified 23-item (with smoking items). For the original 18-item, a total of 420 samples were included in the analysis, which was sufficient for factor analysis as indicated with KMO of 0.82, which is within the recommended range (0.8 to 1). The Bartlett’s Test of Sphericity was significant (X2 = 4235.007, p-value < 0.001) indicating the data is adequate for factor analysis. As a result, a three-factor solution emerged based on the Scree plot (figure 1), accounting 57.4% of the total variance. Factor loading patterns in the present analysis slightly varied from the original subscales. The domains in the original subscales were risk perception, benefit finding and healthy eating intentions. In our analysis, Item 14 *(‘When I eat at least 5 portions of fruit and vegetables a day I am doing something good for the health of my heart’*) showed a better loading to healthy eating intention, which was loaded to benefit finding in the original study (Appendix 1).

**Figure 1.**
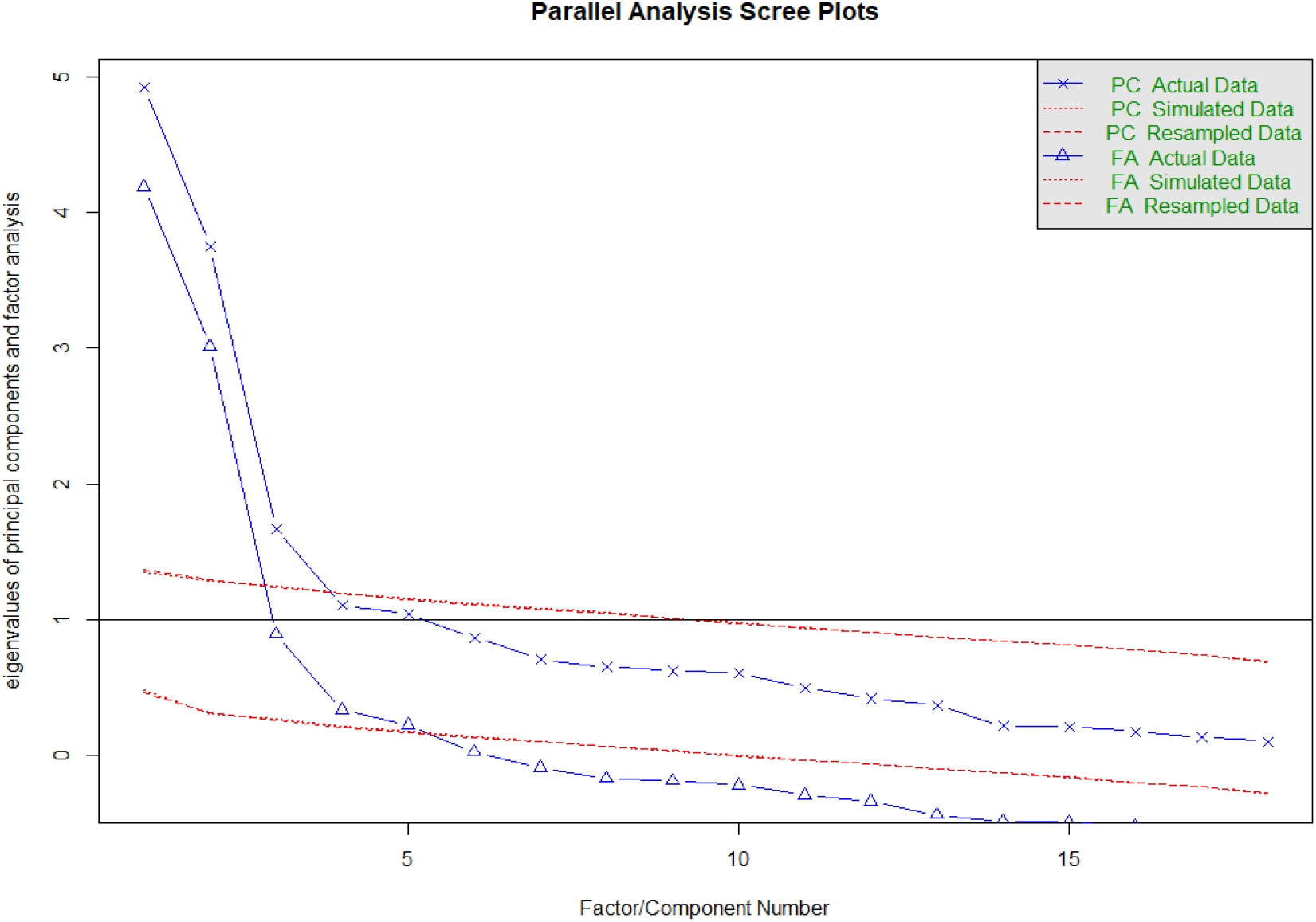
18 item ABCD Questionnaire results from Nottingham dataset.

For the modified 23-item (including the smoking sub-scale), 88 samples were valid and included in the analysis. The KMO was 0.78, which was slightly below the recommended range, but Bartlett’s Test of Sphericity was significant (X2 = 1223.459, p-value < 0.001), indicating adequacy for factor analysis. The analysis showed that the smoking items loaded to another latent construct resulting in four factors in total (figure 2).

**Figure 2.**
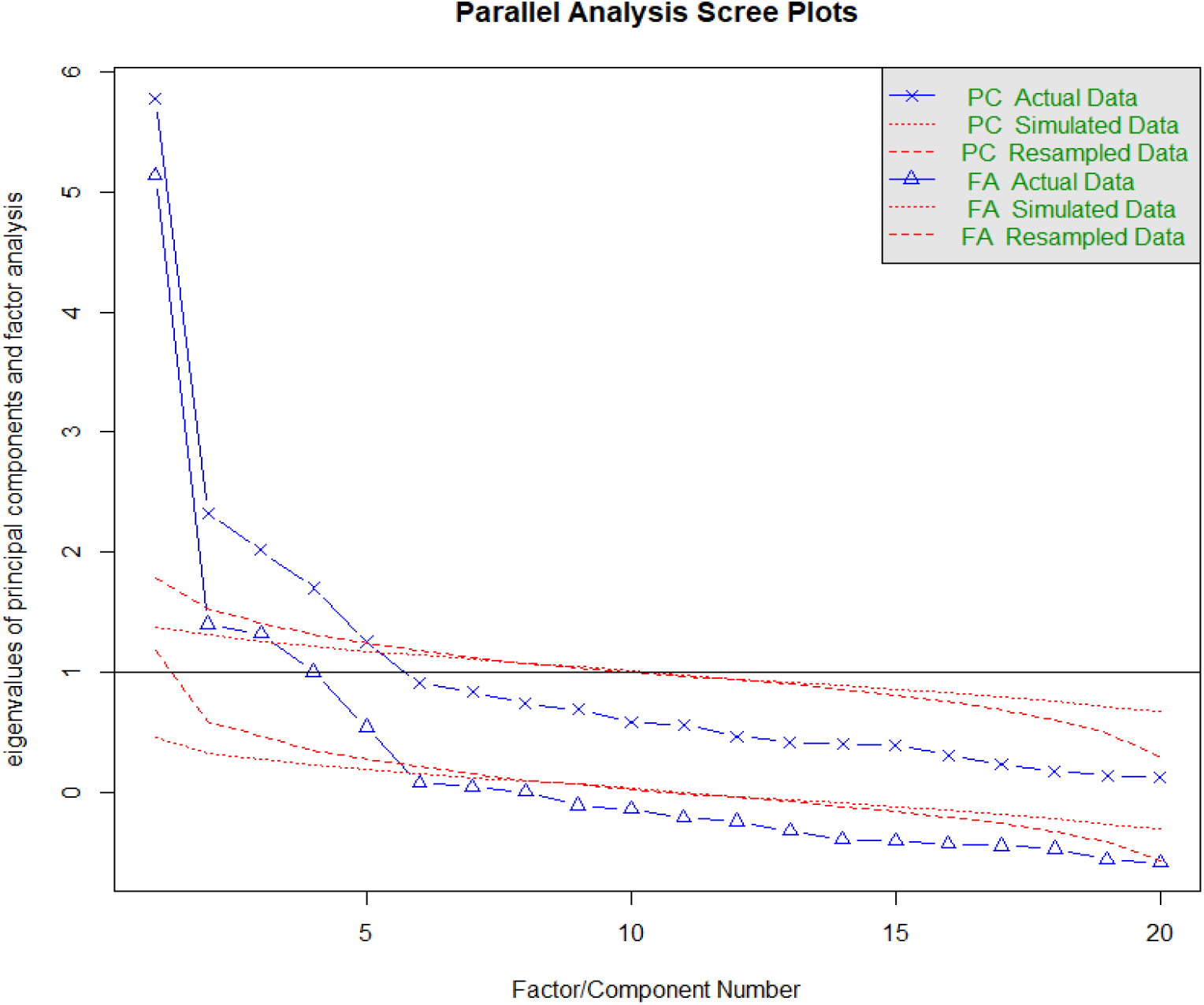
Modified ABCD Questionnaire 20 items with smoking. Nottingham dataset.

### Confirmatory Factor Analysis of the published ABCD Risk Questionnaire

A Confirmatory Factor Analysis was undertaken using the SPICES Nottingham dataset to investigate further. Conducting CFA allowed us to construct the sub-scales of the published ABCD Risk Questionnaire in a three-factor measurement model and test its fit against relevant indices. Original 18 item survey comprising three sub-scales (Perceived Risk of Heart Attack/Stroke 8 items; Perceived Benefits and Intentions to Change 7 items; Healthy Eating Intentions 3 items) were used to create measurement model in SPSS Amos 25. The model was then updated to include an additional 5 item sub-scale relating to smoking behaviours.

### Editing the measurement model

The CFA measurement model was then reconstructed removing items which had confused participants and generated high inter-item correlations, and additionally re-assigning an item relating to dietary behaviour into the dietary behaviour sub-scale. This resulted in a four-factor model (Perceived Risk of Heart Attack/ Stroke’ 6 items; ‘Perceived Benefits and Intentions to Exercise’ 6 items; ‘Healthy Eating Intentions’ 4 items, Perceived Benefits and Intentions to Reduce Smoking’ 5 items).

Analysis properties were set to Estimation: Maximum Likelihood, Fit the saturated and independence models; Outputs: Minimisation history, Standardised estimates, Squared multiple correlations, Residual moments, Modification indices, Factor score weights, Covariances of estimates, Correlations of estimates, Threshold for modification indices =4. Calculated model fit estimates considered: CMIN (Chi square), p, CMIN/DF, RMR, TLI, CFI, RMSEA. Modification Indices considered: Covariances between error terms within sub-scales.

Similarly, in the 23-item factor analysis, item 14 was loaded to the healthy eating intention. The model fit indices showed a slight improvement as indicated in table 3.

**Table 1.**
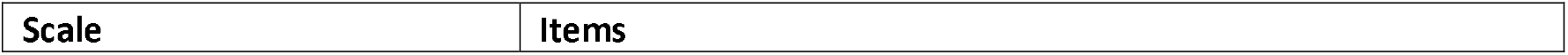
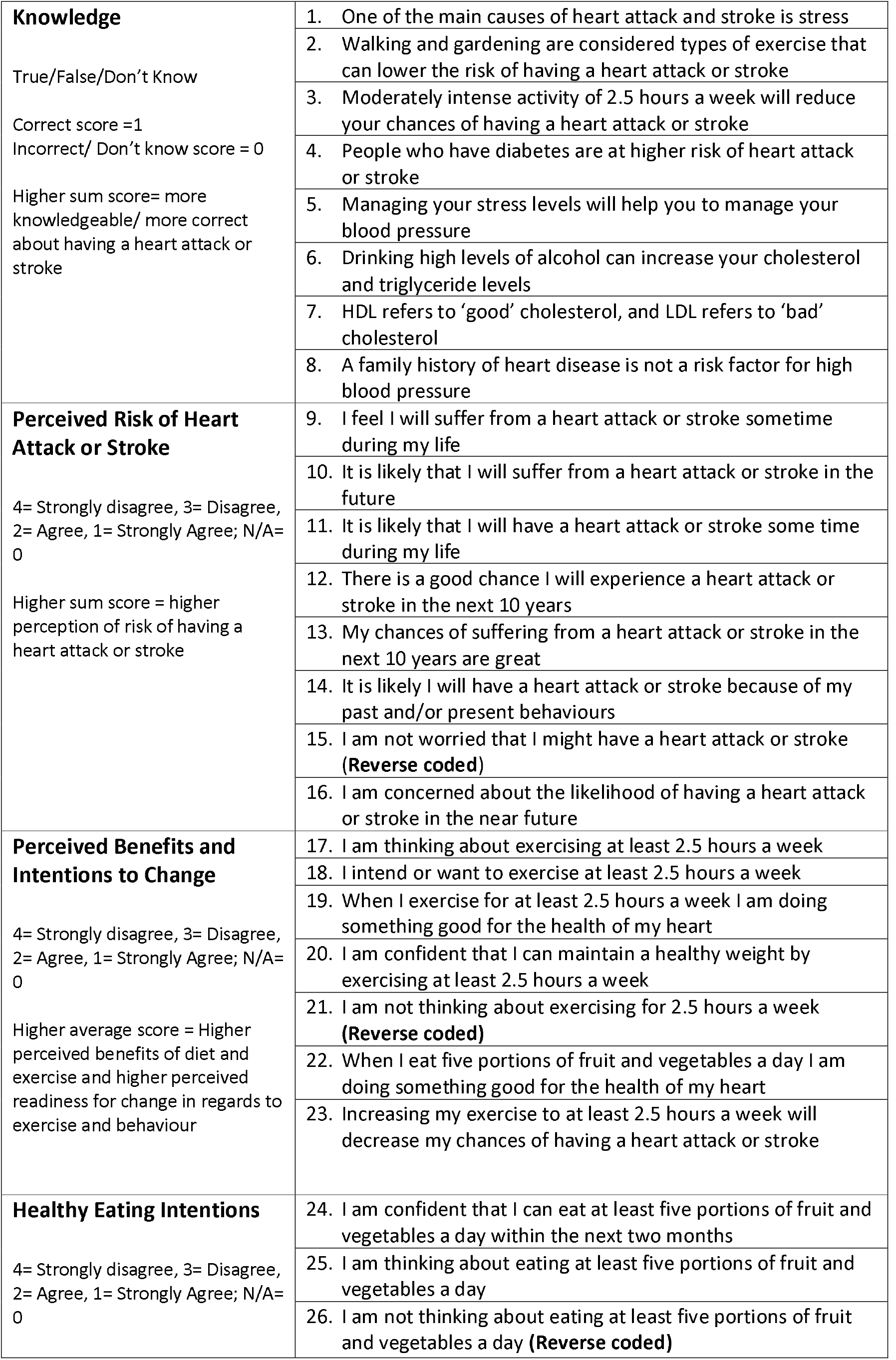
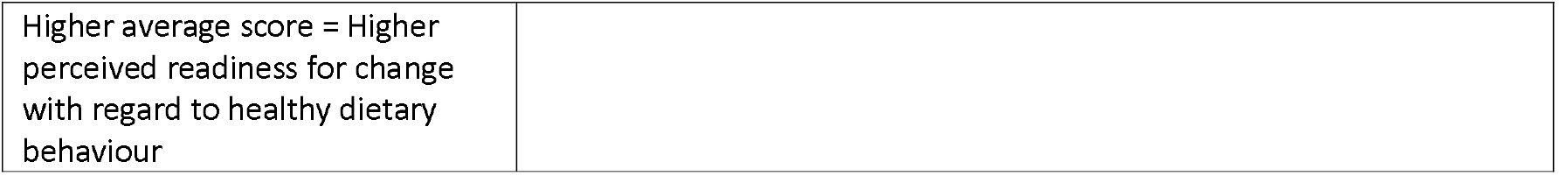
Published ABCD Risk Questionnaire

**Table 2.** Additional ‘smoking’ sub-scale

|  |  |
| --- | --- |
| **Benefits and Intentions to Stop Smoking**<br>4= Strongly disagree, 3= Disagree, 2= Agree, 1= Strongly Agree; N/A= 0<br>Higher average score = Higher perceived readiness for change with regard to healthy dietary behaviour | 27. I am thinking of stopping smoking within two months |
|  | 28. I have reduced or stopped smoking |
|  | 29. I intend or want to stop smoking |
|  | 30. If I stop smoking it will reduce my chances of having a heart attack or stroke |
|  | 31. I am not thinking about stopping smoking |

**Table 3.** CFA fit indices for the original and modified ABCD Questionnaire measurement models

| **Original 18 item ABCD** |  |  |  |  |  |  |
| --- | --- | --- | --- | --- | --- | --- |
| CMIN | P | CMIN/DF | TLI | CFI | RMSEA | RMR |
| 714.941 | .000 | 5.416 | .826 | .850 | .097 | .049 |
| **Updated 23 item ABCD with Smoking sub-scale** |  |  |  |  |  |  |
| CMIN | P | CMIN/DF | TLI | CFI | RMSEA | RMR |
| 994.931 | .000 | 4.442 | .865 | .881 | .086 | .049 |
| **Edited 20 item ABCD with Smoking sub-scale** |  |  |  |  |  |  |
| CMIN | P | CMIN/DF | TLI | CFI | RMSEA | RMR |
| 638.973 | .000 | 3.896 | .881 | .897 | .079 | .052 |
| **Modified 20 item ABCD with Smoking sub-scale** |  |  |  |  |  |  |
| CMIN | P | CMIN/DF | TLI | CFI | RMSEA | RMR |
| 385.312 | .000 | 2.439 | .941 | .951 | .056 | .046 |

Based on factor loading and face validity, we also tested a slightly shorter version of the questionnaire, 20-items including five smoking items and the result shows that the model fit improved (CFI=0.941; TLI=0.951; RMSEA=0.056, SRMR=0.046).

The three published factors achieved a poor fit in CFA (Table 3). Including the five smoking related items which had performed strongly in EFA as their own latent factor improved overall model fit slightly, but not to an acceptable level.

### Modification of the measurement model

Reviewing modification indices and expected parameter changes for factor loadings and measurement intercepts we observed an extreme covariance value (116.812) and parameter change (.209) between two of the risk perception items (‘there is a good chance that I will experience a heart attack or stroke in the next 10 years’ and ‘my chances of suffering a heart attack or stroke in the next 10 years are great’) which had caused confusion for participants in our study.

Removing one of these two items (item #13), and the two other duplicative items (items #9 & #10) from the ‘perceived risk of heart attack or stroke’ sub-scale retains the conceptual spread of risk embodied by the items (lifetime, 10 year, near future, behaviour related). Moving the diet related item (#22) which appears in the ‘perceived benefits and intentions to change’ over to the ‘healthy eating intentions’ sub-scale might allow greater clarity for researchers analysing results from the questionnaire. Co-varying items within sub-scales that generated values above 20 (a high cut-off due to large sample used) resulted in acceptable or good fit across all sub-scales. Each of the three behaviour related sub-scales now contain items drawn from HBM, TTM and SE models providing a sound conceptual basis for comparison. Using EFA to check these results shows the modified sub-scale structure performs better than the published version (all EFA results Appendix 1).

## DISCUSSION

Inadequate knowledge and/or a gap between perceived and actual CVD risk in the population could be an obstacle to better health outcomes. Improving an individual’s CVD knowledge and risk perception may be important in improving a healthy lifestyle. Measuring CVD knowledge and risk perception may be a method to initiate a healthy lifestyle intervention as well as to monitor and evaluate the impact of interventions. Following this rationale, Woringer and colleagues developed the ABCD Risk questionnaire in order to measure CVD knowledge and risk perception. In this study, we re-validated the tool on a sample of the general population in Nottingham to confirm the psychometric properties.

In this Nottingham sample the proportion of current smokers was 15.5% which is lower than the England average (18%), and lower than the Nottingham city sample average (20.6%) based on the ONS Annual Population Survey^xxv^. ONS notes that smoking prevalence estimates by local authority can fluctuate due to smaller sample sizes. Our SPICES Nottingham sample cohort also includes some participants from neighbouring Local Authorities with different recorded rates of smoking.

The 88 participants in this study who reported smoking is a low number for pilot testing of psychometric scales but it does exceed a 10:1 ratio of cases to variables making it reasonable to proceed to analysis.

Based on EFA and CFA, we confirmed a three-factor structure, which is somewhat similar to the original sub-scales. However, in our analysis item 14 *(‘When I eat at least 5 portions of fruit and vegetables a day I am doing something good for the health of my heart’’*) showed a better loading to the ‘healthy eating intentions’ sub-scale, in contrast to the factor loading in the original study, which placed this item in the ‘perceived benefits and intentions to change’ sub-scale. This is the only item which loaded onto a different sub-scale when using the Nottingham dataset, all others continued to load onto their original factors although many of these loaded weakly and failed to meet usual thresholds for validity (Appendix 1). The larger numbers of participants in our dataset (466 compared to 110) provides greater statistical confidence in the reported results, and we therefore adopted this change in the Confirmatory Factor Analysis which also indicated a better fit when item 14 loaded to Healthy Eating Intentions.

These results suggest that the additional five smoking items perform acceptably and should be incorporated into future applications of the ABCD Risk Questionnaire.

### Other observations

Researchers in the Nottingham SPICES team administering the questionnaire during fieldwork reported that three items within the ‘Perception of Risk of Heart Attack/Stroke’ sub-scale caused consistent difficulties for respondents due to apparent duplication and confusion over fine semantic differences. It was difficult for participants to see a semantic difference between statements 9, 10, 11, and 12, 13 respectively. For items 9, 10, and 11, if we agree that *suffer from* and *have* are synonymous, it is hard to differentiate between *in the future and some time during my life* because you would imagine that respondents will be thinking about the future in both cases.

For the questionnaire to be reliable across all sections of the population, including those with limited ability in English (whether native or non-native, first, second or additional language, etc.) who may find it particularly hard to differentiate with any confidence between different pairs/sets of statements with largely synonymous meanings, this confusion is a problem. Items 12 and 13 seem to differ mainly only in the possible interpretation of a difference of degree between good and great.

These face validity issues and their impact can be observed in the inter-item correlation results generated during item reliability analysis. In the original study, two items in the perception of risk sub-scale had been rejected due to correlations in excess of 0.6 leaving 8 items. Of these remaining 8 items half had inter-item correlations which exceeded 0.6 when tested against the Nottingham dataset. These were items 9, 10, 11, and 12 which generated inter-item correlation values of .832, .869, .616, and .729 respectively. Removing items 9, 10, and 13 does not reduce the conceptual range of the ‘perception of risk’ subscale which is framed temporally from immediate threat to lifetime risk, it simply removes the duplicate or confusing items. Testing this shortened scale with factor analysis strengthens both item and scale reliability and improves factor loadings (Appendix 1). We recommend that future versions of the English language ABCD Risk Questionnaire adopt these edits (Appendix 2).

## CONCLUSIONS

The published English language version of the ABCD Risk Questionnaire, with the removal of three problematic ‘perception’ items, the shift of one item from the ‘perceived benefits and intentions to change’ sub-scale into the ‘healthy eating intentions’ sub-scale, and the addition of a 5 item ‘smoking’ sub-scale performs sufficiently well in validity, reliability and factor analysis with an independent, larger sample to confirm the generalisability of its original published findings. This result supports continued use of the ABCD Risk Questionnaire in the field of CVD prevention research and practice. The inclusion of a smoking behaviours sub-scale is likely to increase its relevance where smoking behaviours still account for a large proportion of individually modifiable CVD risk in a target population. Although criterion validity has now been established for the ‘Perception of risk of heart attack/stroke sub-scale’ by two published studies, the utility of the remaining sub-scales individually or in combination has been under-examined. Future studies should investigate the criterion validity of these sub-scales and the conceptual strength of the items and variables from which they have been composed in order to unambiguously position the resulting survey instrument and evaluate its utility in CVD prevention and treatment practices.

## Supporting information

Factor result tables

Modified ABCD Risk Questionnaire

Participant information and consent

Item analysis result tables

Correlations table

## Data Availability

Available to all reasonable requests

