## Supplementary material for "Validating a previously untested ‘Intentions and Beliefs around Smoking’ sub-scale for inclusion in the published ‘Attitudes and Beliefs about Cardiovascular Disease (ABCD) Risk Questionnaire’ using a cross-sectional sample": Factor result tables

**Without smoking items** –

Non-missing samples: 420

Bartlett's Test of Sphericity (X2 = 4235.007, p-value < 0.001)

The overall KMO is 0.82, which is within the recommended range (0.8 to 1).

**EFA results**

- The root mean square of the residuals (RMSR) is 0.05
- Tucker Lewis Index of factoring reliability = 0.77
- RMSEA index = 0.121 and the 90 % confidence intervals are 0.113 0.129
- BIC = 165.35

**Scree plot**

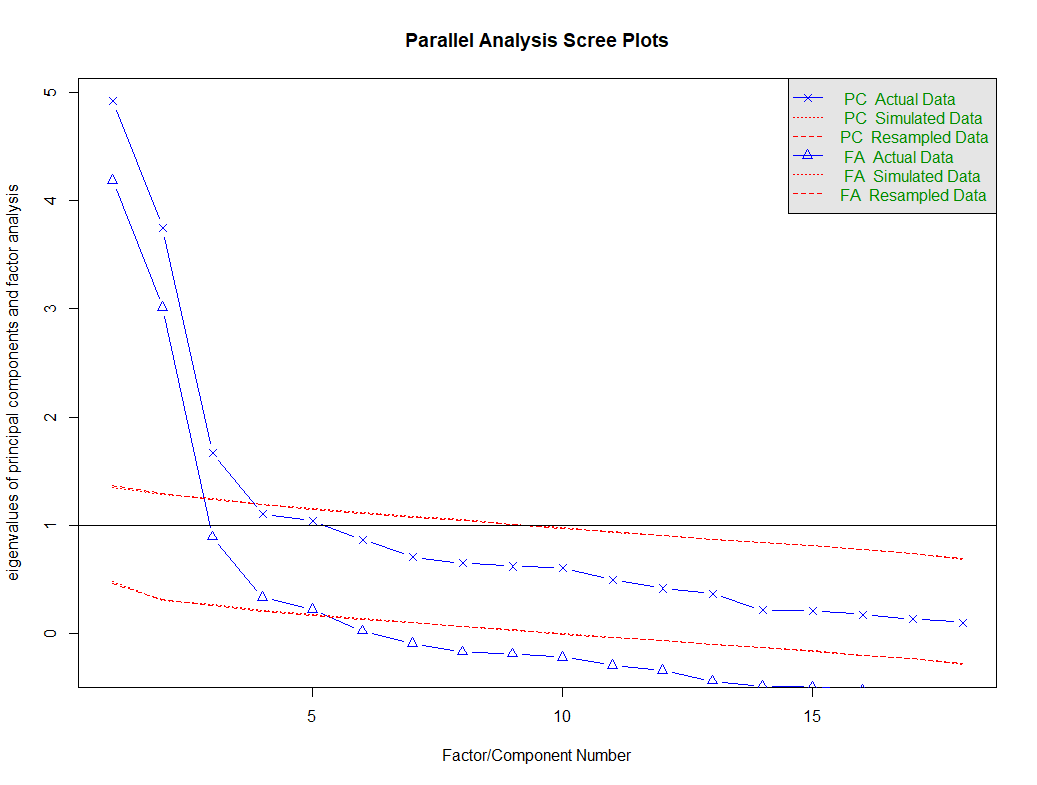

**Factor loadings**

**Table ___. Factor loadings of the exploratory factor analysis of the risk scale without the smoking items**

| Item | Factor2 | Factor1 | Factor3 | communalit | uniqueness |
| --- | --- | --- | --- | --- | --- |
| suffer_heartattack | 0.86 | 0.02 | -0.03 | 0.74 | 0.26 |
| hrtattack_stroke_future | 0.91 | 0.05 | 0.00 | 0.82 | 0.18 |
| attck_stoke_during_life | 0.88 | 0.01 | 0.01 | 0.77 | 0.23 |
| hrtattack_next_10yrs | 0.73 | -0.07 | 0.01 | 0.55 | 0.45 |
| highchance_hrtattck_10yrs | 0.65 | -0.10 | 0.01 | 0.44 | 0.56 |
| hrtattack_past_fut_behav | 0.56 | -0.03 | -0.01 | 0.32 | 0.68 |
| reversenoworry | 0.28 | -0.11 | 0.10 | 0.10 | 0.90 |
| concern_hrtattack | 0.40 | -0.02 | 0.11 | 0.16 | 0.84 |
| think_exercise | -0.02 | 0.87 | -0.06 | 0.73 | 0.27 |
| want_exercise | -0.01 | 0.91 | -0.04 | 0.80 | 0.20 |
| exercise_gud_hrt_hlth | 0.02 | 0.69 | 0.10 | 0.53 | 0.47 |
| confident_hlth_wgt | -0.05 | 0.45 | 0.19 | 0.31 | 0.69 |
| revnotthinkPA | 0.04 | 0.56 | 0.05 | 0.34 | 0.66 |
| fruit_veg_gud_hrthlth | 0.02 | 0.37 | 0.35 | 0.36 | 0.64 |
| high_exerc_low_hrtattack | 0.02 | 0.39 | 0.27 | 0.30 | 0.70 |
| diet_1 | -0.04 | 0.07 | 0.64 | 0.46 | 0.54 |
| diet_2 | 0.01 | -0.01 | 0.93 | 0.85 | 0.15 |
| revdiet3 | -0.01 | -0.03 | 0.78 | 0.60 | 0.40 |

|  | Factor 2 | Factor 1 | Factor 3 |
| --- | --- | --- | --- |
| SS loadings | 3.86 | 3.04 | 2.28 |
| Proportion Var | 0.21 | 0.17 | 0.13 |
| Cumulative Var | 0.21 | 0.38 | 0.51 |
| Proportion Explained | 0.42 | 0.33 | 0.25 |
| Cumulative Proportion | 0.42 | 0.75 | 1.00 |

**With smoking item**

Non-missing samples: 88

The overall KMO is 0.78, which is slightly below the recommended range (0.8 to 1).

The Bartlett’s test of Sphericity is significant (X2 = 1223.459, p-value < 0.001), indicating the sample adequacy for factor analysis.

**EFA results**

- The root mean square of the residuals (RMSR) is 0.06
- Tucker Lewis Index of factoring reliability = 0.69
- RMSEA index = 0.129 and the 90 % confidence intervals are 0.124 and 0.136
- BIC = 440.9

**Scree plot**

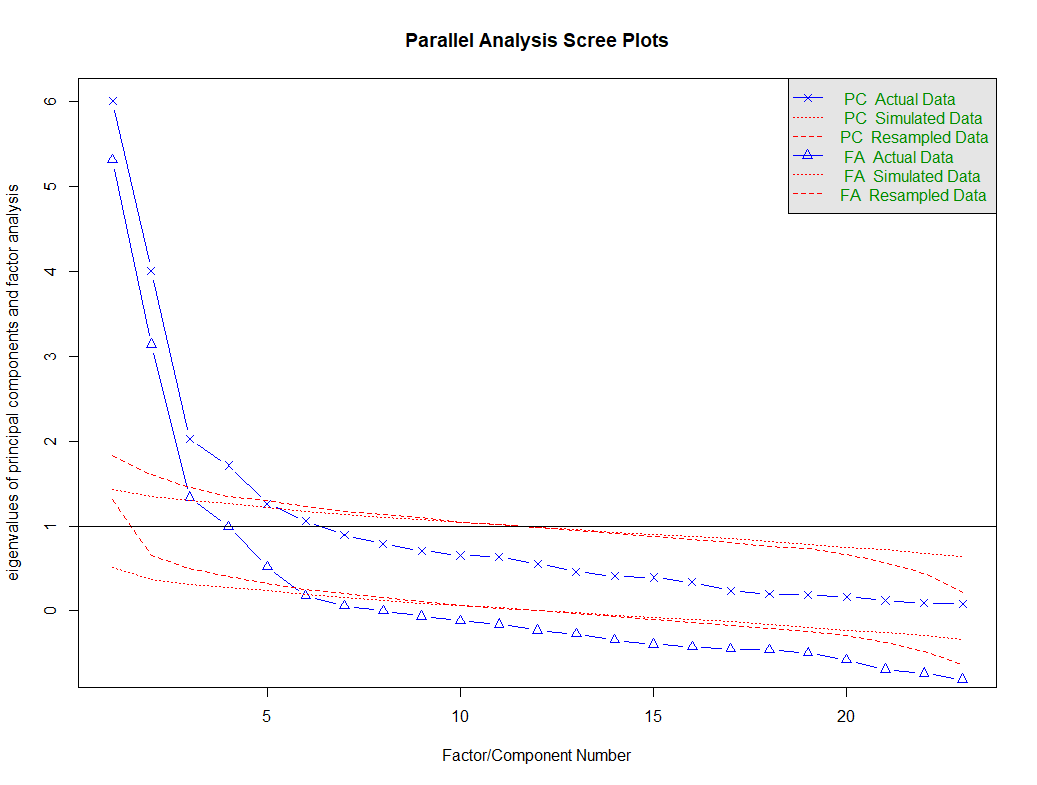

Factor loadings

**Table ___. Factor loadings of the exploratory factor analysis of the ABCD Questionnaire with the smoking items**

| Item | Factor2 | Factor3 | Factor1 | Factor4 | Communality | Uniqueness |
| --- | --- | --- | --- | --- | --- | --- |
| suffer_heartattack | 0.86 | -0.1 | 0.05 | -0.02 | 0.76 | 0.24 |
| hrtattack_stroke_future | 0.91 | 0.06 | 0.02 | -0.01 | 0.82 | 0.18 |
| attck_stoke_during_life | 0.88 | 0.02 | 0 | 0 | 0.77 | 0.23 |
| hrtattack_next_10yrs | 0.72 | 0 | -0.09 | 0.01 | 0.54 | 0.46 |
| highchance_hrtattck_10yrs | 0.64 | -0.03 | -0.1 | 0.01 | 0.45 | 0.55 |
| hrtattack_past_fut_behav | 0.57 | -0.07 | 0 | 0 | 0.33 | 0.67 |
| reversenoworry | 0.28 | 0.02 | -0.14 | 0.1 | 0.1 | 0.9 |
| concern_hrtattack | 0.41 | 0.19 | -0.12 | 0.08 | 0.19 | 0.81 |
| think_exercise | -0.03 | -0.05 | 0.88 | -0.02 | 0.73 | 0.27 |
| want_exercise | -0.02 | 0.05 | 0.87 | -0.02 | 0.79 | 0.21 |
| exercise_gud_hrt_hlth | 0.03 | 0.17 | 0.62 | 0.09 | 0.55 | 0.45 |
| confident_hlth_wgt | -0.05 | 0.09 | 0.42 | 0.18 | 0.32 | 0.68 |
| revnotthinkPA | 0.02 | 0 | 0.53 | 0.09 | 0.33 | 0.67 |
| fruit_veg_gud_hrthlth | 0.04 | 0.07 | 0.35 | 0.35 | 0.36 | 0.64 |
| high_exerc_low_hrtattack | 0.04 | 0.12 | 0.37 | 0.24 | 0.32 | 0.68 |
| diet_1 | -0.04 | -0.05 | 0.12 | 0.64 | 0.45 | 0.55 |
| diet_2 | 0.01 | 0 | 0.02 | 0.89 | 0.8 | 0.2 |
| revdiet3 | -0.01 | 0 | -0.06 | 0.83 | 0.66 | 0.34 |
| smoking_1 | 0.06 | 0.78 | 0.12 | -0.06 | 0.67 | 0.33 |
| smoking_2 | -0.03 | 0.83 | 0.02 | -0.01 | 0.71 | 0.29 |
| smoking_3 | -0.05 | 0.9 | -0.02 | -0.01 | 0.8 | 0.2 |
| smoking_4 | 0.16 | 0.58 | 0.09 | 0.08 | 0.43 | 0.57 |
| revsmoke5 | -0.12 | 0.56 | -0.2 | 0.17 | 0.35 | 0.65 |

|  | Factor 2 | Factor 3 | Factor 1 | Factor 4 |
| --- | --- | --- | --- | --- |
| SS loadings | 3.90 | 3.00 | 2.97 | 2.33 |
| Proportion Var | 0.17 | 0.13 | 0.13 | 0.10 |
| Cumulative Var | 0.17 | 0.30 | 0.43 | 0.53 |
| Proportion Explained | 0.32 | 0.25 | 0.24 | 0.19 |
| Cumulative Proportion | 0.32 | 0.57 | 0.81 | 1.00 |

**Modified scale (20-items including the smoking items)**

Non-missing samples: 89

The overall KMO is 0.79, which is slightly below the recommended range (0.8 to 1).

The Bartlett’s test of Sphericity is significant (X2 = 915.41, p-value < 0.001), indicating the sample adequacy for factor analysis.

**EFA results**

- The root mean square of the residuals (RMSR) is 0.06
- Tucker Lewis Index of factoring reliability = 0.72
- RMSEA index = 0.118 and the 90 % confidence intervals are 0.111 and 0.126
- BIC = 153.72

**Scree plot**

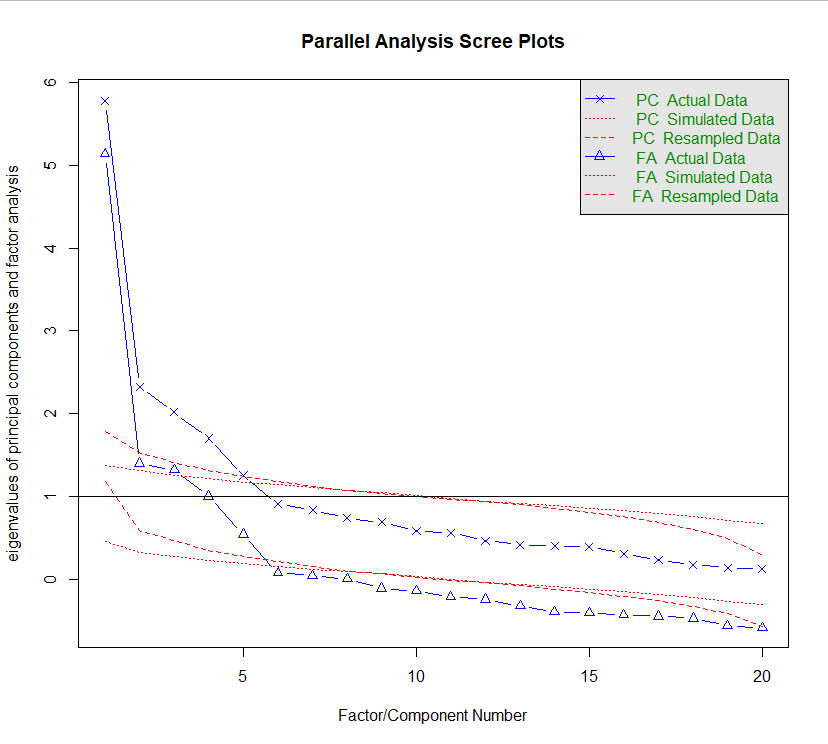

**Table ___. Factor loadings of the exploratory factor analysis of the modified ABCD questionnaire (20 items including the smoking items)**

| Item | Factor3 | Factor1 | Factor4 | Factor2 | Communality | Uniqueness |
| --- | --- | --- | --- | --- | --- | --- |
| suffer_heartattack | -0.08 | 0.04 | -0.03 | 0.76 | 0.60 | 0.40 |
| hrtattack_next_10yrs | 0.02 | -0.08 | -0.01 | 0.68 | 0.48 | 0.52 |
| hrtattack_past_fut_behav | -0.04 | 0.01 | -0.01 | 0.61 | 0.38 | 0.62 |
| reversenoworry | 0.04 | -0.13 | 0.10 | 0.35 | 0.14 | 0.86 |
| concern_hrtattack | 0.22 | -0.11 | 0.07 | 0.45 | 0.23 | 0.77 |
| think_exercise | -0.06 | 0.88 | -0.02 | -0.04 | 0.74 | 0.26 |
| want_exercise | 0.05 | 0.87 | -0.02 | -0.02 | 0.79 | 0.21 |
| exercise_gud_hrt_hlth | 0.17 | 0.62 | 0.09 | 0.04 | 0.55 | 0.45 |
| confident_hlth_wgt | 0.09 | 0.42 | 0.18 | -0.06 | 0.32 | 0.68 |
| revnotthinkPA | 0.01 | 0.53 | 0.09 | 0.03 | 0.32 | 0.68 |
| fruit_veg_gud_hrthlth | 0.08 | 0.35 | 0.35 | 0.07 | 0.37 | 0.63 |
| high_exerc_low_hrtattack | 0.13 | 0.37 | 0.24 | 0.06 | 0.32 | 0.68 |
| diet_1 | -0.06 | 0.12 | 0.64 | -0.05 | 0.46 | 0.54 |
| diet_2 | 0.00 | 0.02 | 0.89 | 0.01 | 0.80 | 0.20 |
| revdiet3 | 0.00 | -0.06 | 0.83 | -0.01 | 0.67 | 0.33 |
| smoking_1 | 0.78 | 0.12 | -0.06 | 0.04 | 0.66 | 0.34 |
| smoking_2 | 0.83 | 0.02 | -0.01 | -0.03 | 0.70 | 0.30 |
| smoking_3 | 0.89 | -0.02 | -0.01 | -0.07 | 0.80 | 0.20 |
| smoking_4 | 0.59 | 0.10 | 0.07 | 0.18 | 0.43 | 0.57 |
| revsmoke5 | 0.56 | -0.20 | 0.17 | -0.10 | 0.34 | 0.66 |

|  | Factor3 | Factor1 | Factor4 | Factor2 |
| --- | --- | --- | --- | --- |
| SS loadings | 3.00 | 2.96 | 2.33 | 1.80 |
| Proportion Var | 0.15 | 0.15 | 0.12 | 0.09 |
| Cumulative Var | 0.15 | 0.30 | 0.41 | 0.50 |
| Proportion Explained | 0.30 | 0.29 | 0.23 | 0.18 |
| Cumulative Proportion | 0.30 | 0.59 | 0.82 | 1.00 |
