## Supplementary material for "Validating a previously untested ‘Intentions and Beliefs around Smoking’ sub-scale for inclusion in the published ‘Attitudes and Beliefs about Cardiovascular Disease (ABCD) Risk Questionnaire’ using a cross-sectional sample": Modified ABCD Risk Questionnaire

| **Scale** | **Items** | **Coding** |
| --- | --- | --- |
| **Perceived Risk of Heart Attack or Stroke** | 1. It is likely that I will have a heart attack or stroke sometime in my life | 4= Strongly disagree, 3= Disagree, 2= Agree, 1= Strongly Agree; N/A= 0 |
|  | 2. There is a good chance I will experience a heart attack or stroke in the next 10 years | 4= Strongly disagree, 3= Disagree, 2= Agree, 1= Strongly Agree; N/A= 0 |
|  | 3. It is (more) likely I will have a heart attack or stroke because of my past and/or present behaviours | 4= Strongly disagree, 3= Disagree, 2= Agree, 1= Strongly Agree; N/A= 0 |
|  | 4. I am not worried that I might have a heart attack or stroke | **REVERSE CODED**  4= Strongly disagree, 3= Disagree, 2= Agree, 1= Strongly Agree; N/A= 0 |
|  | 5. I am concerned about the likelihood of having a heart attack or stroke in the near future | 4= Strongly disagree, 3= Disagree, 2= Agree, 1= Strongly Agree; N/A= 0 |
| **Perceived Benefits and Intentions to Exercise** | 6. I am thinking about exercising at least 2.5 hours a week | 4= Strongly disagree, 3= Disagree, 2= Agree, 1= Strongly Agree; N/A= 0 |
|  | 7. I intend or want to exercise at least 2.5 hours a week | 4= Strongly disagree, 3= Disagree, 2= Agree, 1= Strongly Agree; N/A= 0 |
|  | 8. When I exercise for at least 2.5 hours a week I am doing something good for the health of my heart | 4= Strongly disagree, 3= Disagree, 2= Agree, 1= Strongly Agree; N/A= 0 |
|  | 9. I am confident that I can maintain a healthy weight by exercising at least 2.5 hours a week | 4= Strongly disagree, 3= Disagree, 2= Agree, 1= Strongly Agree; N/A= 0 |
|  | 10. I am not thinking about exercising for 2.5 hours a week | **REVERSE CODED**  4= Strongly disagree, 3= Disagree, 2= Agree, 1= Strongly Agree; N/A= 0 |
|  | 11. Increasing my exercise to at least 2.5 hours a week will decrease my chances of having a heart attack or stroke | 4= Strongly disagree, 3= Disagree, 2= Agree, 1= Strongly Agree; N/A= 0 |
| **Perceived Benefit and Healthy Eating Intentions** | 12. I am confident that I can eat at least five portions of fruit and vegetables a day within the next two months | 4= Strongly disagree, 3= Disagree, 2= Agree, 1= Strongly Agree; N/A= 0 |
|  | 13. I am thinking about eating at least five portions of fruit and vegetables a day | 4= Strongly disagree, 3= Disagree, 2= Agree, 1= Strongly Agree; N/A= 0 |
|  | 14. I am not thinking about eating at least five portions of fruit and vegetables a day | **REVERSE CODED**  4= Strongly disagree, 3= Disagree, 2= Agree, 1= Strongly Agree; N/A= 0 |
|  | 15. When I eat five portions of fruit and vegetables a day I am doing something good for the health of my heart | 4= Strongly disagree, 3= Disagree, 2= Agree, 1= Strongly Agree; N/A= 0 |
| **Benefits and Intentions to Stop Smoking** | 16. I am thinking of stopping smoking within two months | 4= Strongly disagree, 3= Disagree, 2= Agree, 1= Strongly Agree; N/A= 0 |
|  | 17. I have reduced or stopped smoking | 4= Strongly disagree, 3= Disagree, 2= Agree, 1= Strongly Agree; N/A= 0 |
|  | 18. I intend or want to stop smoking | 4= Strongly disagree, 3= Disagree, 2= Agree, 1= Strongly Agree; N/A= 0 |
|  | 19. If I stop smoking it will reduce my chances of having a heart attack or stroke | 4= Strongly disagree, 3= Disagree, 2= Agree, 1= Strongly Agree; N/A= 0 |
|  | 20. I am not thinking about stopping smoking | **REVERSE CODED**  4= Strongly disagree, 3= Disagree, 2= Agree, 1= Strongly Agree; N/A= 0 |
