## Supplementary material for "Validating a previously untested ‘Intentions and Beliefs around Smoking’ sub-scale for inclusion in the published ‘Attitudes and Beliefs about Cardiovascular Disease (ABCD) Risk Questionnaire’ using a cross-sectional sample": Participant information and consent

**‘SPICES’ Heart Diseases Prevention Research**

Introduction to SPICES research

Nottingham Trent University is part of an international research team investigating ways to build good practice in the prevention of Heart Diseases. Researchers and doctors have a lot of evidence about what causes heart diseases and what prevents them. Heart Diseases are now the biggest cause of death globally, and one of the leading causes of disability, so the more people know what the doctors know, the better they can protect themselves and maintain a good quality of life.

The research project is called 'SPICES' and here in Nottingham we are going to see if working with people in the community instead of at the doctor's surgery, we can spread the message quicker and further.

If you choose to take part we will ask you to complete a simple survey. From the we will be able see how well you are looking after your heart in terms of your lifestyle. Then there will be three possible options:

If the data you provide suggests you may need to make some lifestyle changes we will recommend that you make an appointment to see your doctor. As researchers we cannot give any medical advice, but it would be inappropriate for us to ignore any signs of an unhealthy lifestyle that could give rise to heart problems.

If the data you provide suggests you have a healthy lifestyle, then this is positive news and we'll talk to you about how you might be able to help the project in other ways.

If you are somewhere in the middle we will show you some simple ways to reduce your risk and stay healthier for longer.

N.B. In all cases, the data you provided is for research purposes only and a decision about your health cannot be made on the basis of questionnaires only. Whilst we advise you to see a doctor if figures are high, lower figures should not be taken to indicate a healthy heart, and the results should not be used to replace medical assessments and the taking of medical advice about other health monitoring strategies. The dividing of participants into three groups is for research purposes only and is not a medical intervention.

If you're interested please complete our survey (It might take about 10 minutes, and you will need a tape measure for one of the questions).

Our researchers will then get in touch with you about ways that we can support you to make your heart healthier. Any information we collect will be kept securely and not shared outside of the research team. Your name and personal details will not be used in any reports, and all our records will be destroyed at the end of the project in line with the relevant GDPR legislation. Additionally you may withdraw your data at any time up to but no later than December 31st 2020 by contacting Mark Bowyer, SPICES Coordinator, Nottingham Trent University 0115 8485574

OK? Let's start with your agreement to take part.

**CONSENT FORM**

**‘SPICES’ Heart Diseases Prevention Research**

You are making a decision to take part. By ticking ALL statements and signing your name below you will indicate that you have read the information provided above and decided to participate.

If you choose to discontinue participation in this study, you may withdraw at any time without judgement, or effect on your status.

| **CONSENT STATEMENT** | | **Please tick if you agree** |
| --- | --- | --- |
| **1.** | I have received, read and understood the SPICES participant information sheet |  |
| **2.** | I am aware that I can withdraw my participation at any time without prejudice, judgement or effect on my status in relation to Nottingham Trent University or its research partners |  |
| **3.** | I understand that information I provide during my participation can be deleted at my request up to but no later than December 31^st^ 2020 |  |
| **4.** | I agree to be contacted by SPICES researchers using the details that I have supplied below |  |
| **5.** | I understand that the collection of data is not part of medical assessment or diagnosis and cannot be relied upon to reach conclusions as to the state of my health |  |
| **5.** | I understand that any information I provide as part of the SPICES research will be managed in accordance with the EU General Data Protection Regulation (GDPR) framework (see SPICES participant information sheet) |  |
| **6.** | I agree to take part in this research project |  |

**Name:**

**Preferred contact details:**

**D.O.B.**

**Gender:**

**Postcode:**

**Signature:**

**Date:**

……………………………………………………………………..

**Staff signature:**

**Date:**
