## Supplementary material for "Validating a previously untested ‘Intentions and Beliefs around Smoking’ sub-scale for inclusion in the published ‘Attitudes and Beliefs about Cardiovascular Disease (ABCD) Risk Questionnaire’ using a cross-sectional sample": Item analysis result tables

Appendix 5. Item Analysis of published ABCD Risk Questionnaire sub-scales plus 5 unpublished items relating to smoking.

| **Perceived Risk of Heart Attack/ Stroke**  **8 Items**  **Cronbach’s Alpha .861** | **Inter-item correlation** | **Corrected Item-total correlation** | **Cronbach’s alpha if item deleted** |
| --- | --- | --- | --- |
| It is likely that I will suffer from a heart attack or stroke in the future | **.832** | .756 | .826 |
| It is likely that I will have a heart attack or stroke some time during my life | **.869** | .777 | .824 |
| I feel I will suffer a heart attack or stroke some time during my life | **.616** | .784 | .824 |
| There is a good chance I will experience a heart attack or stroke in the next 10 years | **.729** | .722 | .832 |
| I am not worried that I might have a heart attack or stroke | .403 | .624 | .843 |
| My chances of suffering a heart attack or stroke in the next 10 years are great | .245 | .544 | .852 |
| It is likely that I will have a heart attack or stroke because of my past/present behaviours | .266 | .319 | **.876** |
| I am concerned about the likelihood of having a heart attack or stroke in the near future | .259 | .387 | **.870** |
| **Perceived Benefits and Intentions to Change**  **7 items**  **Cronbach’s Alpha .801** | **Inter-item correlation** | **Corrected Item-total correlation** | **Cronbach’s alpha if item deleted** |
| I am thinking about exercising at least 2.5 hours a week | **.727** | **.605** | .760 |
| I intend or want to exercise at least 2.5 hours a week | .442 | **.651** | .752 |
| When I exercise for at least 2.5 hours a week I am doing something good for the health of my heart | .426 | .593 | .769 |
| I am confident that I can maintain a healthy weight by exercising at least 2.5 hours a week within the next 2 months | .294 | .452 | .790 |
| I am not thinking about exercising at least 2.5 hours a week | .264 | .508 | .781 |
| When I eat at least 5 portions of fruit and vegetables a day I am doing something good for the health of my heart | .483 | .483 | .783 |
| Increasing my exercise to at least 2.5 hours a week will decrease my chances of having a heart attack or stroke | .326 | .474 | .786 |
| **Healthy Eating Intentions**  **3 items**  **Cronbach’s Alpha .787** | **Inter-item correlation** | **Corrected Item-total correlation** | **Cronbach’s alpha if item deleted** |
| I am confident that I can eat at least 5 portions of fruit and vegetables a day within the next 2 months | .555 | .533 | **.812** |
| I am thinking about eating at least 5 portions of fruit and vegetables a day | .**683** | **.732** | .596 |
| I am not thinking about eating at least 5 portions of fruit and vegetables a day | .424 | **.624** | .713 |
| **Perceived Benefits and Intentions to Stop Smoking**  **5 Items**  **Cronbach’s Alpha .943** | **Inter-item correlation** | **Corrected item-total correlation** | **Cronbach’s alpha if item deleted** |
| I am thinking of stopping smoking within the next 2 months | **.654** | .848 | .932 |
| I have reduced or stopped smoking | **.694** | .751 | **.949** |
| I intend or want to stop smoking | **.829** | .906 | .919 |
| If I stop smoking it will reduce my chances of having a heart attack or stroke | **.834** | .886 | .922 |
| I am not thinking about stopping smoking | **.789** | .872 | .925 |
