## Supplementary material for "Validating a previously untested ‘Intentions and Beliefs around Smoking’ sub-scale for inclusion in the published ‘Attitudes and Beliefs about Cardiovascular Disease (ABCD) Risk Questionnaire’ using a cross-sectional sample": Correlations table

**Appendix 6**

ABCD subscale and selected INTERHEART variable correlation values from Nottingham study compared with values reported in the original Woringer study.

|  |  | Knowledge | Perceived Risk | Perceived Benefit | Healthy Intentions | IMD2010 Quintile | BMI/W2Hr | Qrisk2/  INTERHEART |
| --- | --- | --- | --- | --- | --- | --- | --- | --- |
| Knowledge | Correlation Coefficient |  | -.124**/**  **.013** | -.148**/**  **-.021** | -.106**/**  **-.039** | -.002/  .085 | -.225/  -.084 | -.007/  -.018 |
|  | Sig 2 tailed |  | .236/  .722 | .175/  .645 | .319/  .400 | .986/  .066 | .021/  .082 | .941/  .714 |
|  | N |  | 93/462 | 86/462 | 91/462 | 99/466 | 105/433 | 104/436 |
| Perceived Risk | Correlation Coefficient |  |  | -.195/  -.112 | -.188/  -0.36 | .239/  .039 | .389/  .182 | .220/  .356 |
|  | Sig 2 tailed |  |  | .080/  .016 | .088/  .441 | .025/  .397 | .000/  .000 | .036/  .000 |
|  | N |  |  | 82/462 | 84/462 | 87/466 | 92/433 | 91/436 |
| Perceived Benefits | Correlation Coefficient |  |  |  | .533/  .383 | -.287/  .071 | -.068/  .000 | -.118/  -.164 |
|  | Sig 2 tailed |  |  |  | .000/  .000 | .009/  .127 | .538/  .997 | .284/  .001 |
|  | N |  |  |  | 83/462 | 81/466 | 85/433 | 84/436 |
| Healthy Intentions | Correlation Coefficient |  |  |  |  | -.261/  .098 | .084/  .044 | -.072/  -.079 |
|  | Sig 2 tailed |  |  |  |  | .016/  .034 | .430/  .365 | .504/  .100 |
|  | N |  |  |  |  | 85/466 | 90/462 | 89/436 |
